# Multimodal Data Acquisition at SARS-CoV-2 Drive Through Screening Centers: Setup Description and Experiences in Saarland, Germany

**DOI:** 10.1101/2020.12.08.20240382

**Authors:** Philipp Flotho, Mayur J. Bhamborae, Tobias Grün, Carlos Trenado, David Thinnes, Dominik Limbach, Daniel J. Strauss

## Abstract

SARS-CoV-2 drive through screening centers (DTSC) have been implemented worldwide as a fast and secure way of mass screening. We use DTSCs as a platform for the acquisition of multimodal datasets that are needed for the development of remote screening methods. Our acquisition setup consists of an array of thermal, infrared and RGB cameras as well as microphones and we apply methods from computer vision and computer audition for the contactless estimation of physiological parameters. We have recorded a multimodal dataset of DTSC participants in Germany for the development of remote screening methods and symptom identification. Acquisition in the early stages of a pandemic and in regions with high infection rates can facilitate and speed up the identification of infection specific symptoms and large scale data acquisition at DTSC is possible without disturbing the flow of operation.

## I. Introduction

Research on digital technologies to combat the COVID-19 pandemic includes the computational analysis of video and audio data [1], [2], [3]. Due to their contactless nature, such methods are particularly promising and needed for mass screening purposes [3] and besides fever, there are other atypical and non-severe symptoms (e.g. [4], [5], [6], [7]) which allow for non-contact medical assessment. The value of such screening systems would of course be directly related to the achievable sensitivity and specificity for detecting SARS-CoV-2 infections. However, it is challenging to acquire homogeneous data sets for the development and assessment of such remote systems without interfering with medical services due to the pressure of the ongoing pandemic.

For the rapid collection of samples for polymerase chain reaction (PCR) based screening, drive-through screening centers (DTSCs), e.g., Kwon et al. [8], are already in use in several countries. There are advantages of DTSCs for the acquisition of contactless recordings of the patients: They simulate a scenario, where contactless screening of infectious diseases might be implemented one day, such as the entrance of employee parking area. The exposure of equipment and personnel to patients is minimized and at the same time the exposure of healthy participants to contaminated air or equipment is fully controlled and can be completely avoided as the patients stay seated in their own car.

We propose an acquisition system along with a processing pipeline for rapidly acquiring such data at DTSC without disturbing their flow of medical operation and present a multimodal dataset of DTSC users as well as preliminary evaluations.

## II. Materials and Methods

We recorded our dataset between May and July 2020 at the SARS-CoV-2 DTSC located at the former fairground area in Saarbrücken, State of Saarland, Germany. The study was approved by the responsible ethics committee (ethics commission at the Ärztekammer des Saarlandes, ID No 90/20) and after a detailed explanation of the procedure, all included participants signed a consent form. Admission to and recommendation for the tests was given by the participants’ general practitioner if a patient had a potential SARS-CoV-2 infection based on the Robert Koch Institute’s guidelines [9]. The PCR-test result for SARS-CoV-2 from the individual nose and throat swap was accessible for us at the responsible public health office. The recordings were done through an opened window with the participants sitting in their car. Our multimodal setup consisted of RGB, NIR, depth and thermal cameras as well as microphones (see Fig. 1). We recorded at 120fps (face closeups, RGB), 50fps (thermal camera), 30fps (NIR) and 10fps (high resolution RGB, stereo) and used custom acquisition routines and frame grabbers to minimize the user interaction with the recording systems. The investigators followed the same guidelines for the personal protective equipment (PPE) as the physicians taking the swap samples. Participants waiting for the experiment were regularly informed about the estimated waiting period and had the option to quit the experiment between two recordings, to reduce the contamination with ambient sound during audio recordings. The experiment began with a set of yes/no/unknown questions with the goal to generate uniform voice samples that can be compared between subjects. These also provided additional medical history. The second segment was a free speech sample, where the participants were asked to tell the circumstances that led to them visiting the DTSC. Subsequently, we asked the participants to briefly present their hands from both sides towards the camera, to catch potential cues to skin rashes. In the final segment, participants were asked to take 10 deep breaths and to breathe normally for 30s afterwards. The entire study/data acquisition took around 6 minutes. A total of 436 participants with signed consent form participated in our study, aged 19—86 (mean age 45.6 ± 15.2, 215 males, 221 females, 7 participants did not provide their age). 34 subjects reported chronical or acute respiratory diseases or symptoms (see Fig. 3). Despite a relatively high participation of 36% of the DTSC users in our study, our data set contained only two subjects with SARS-CoV-2 positive PCR results from the swab tests at the DTSC, see discussion.

**Fig. 1.**
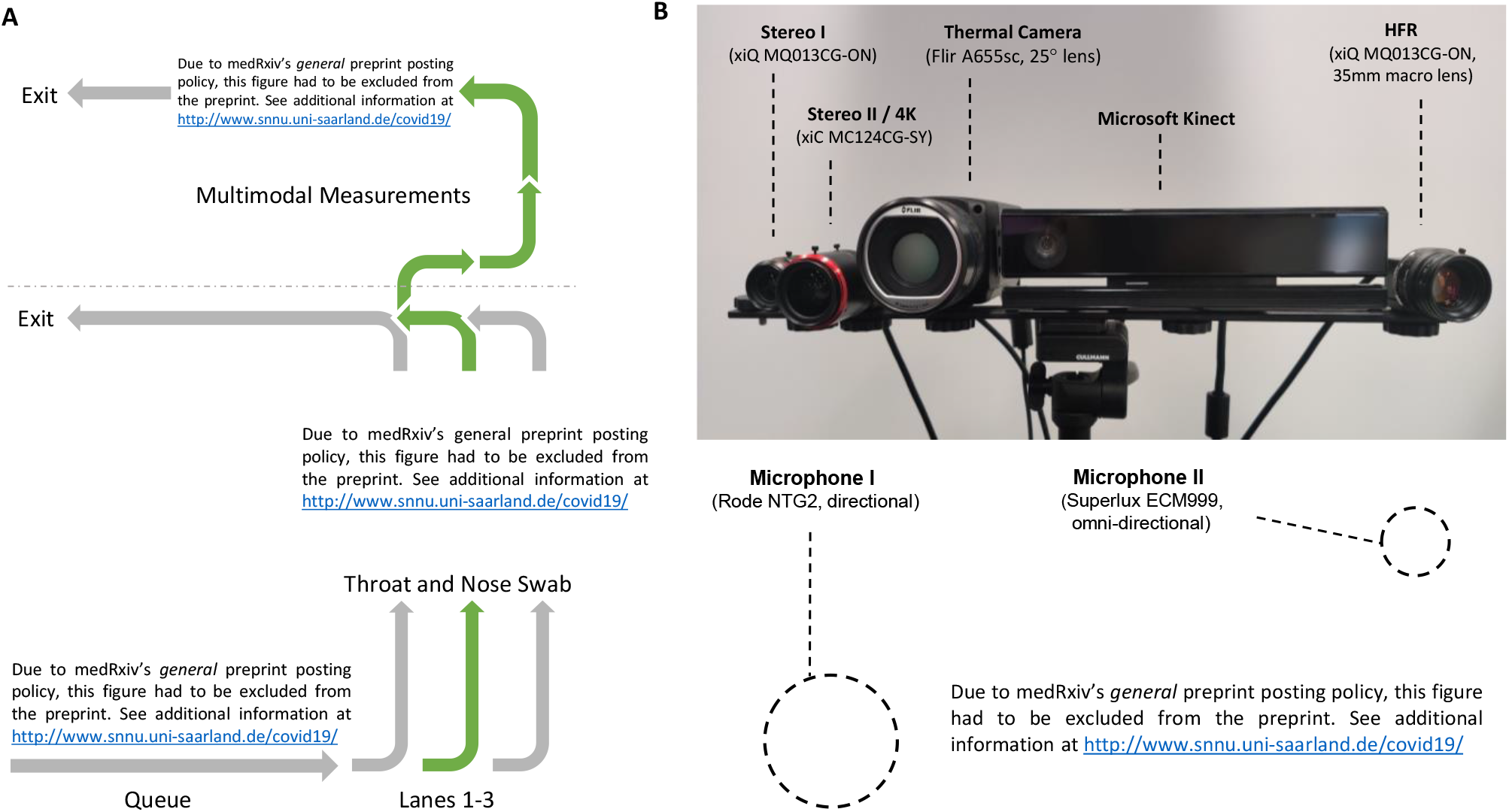
Schematic depiction of the flow at the SARS-CoV-2 DTSC (A) and of the multimodal acquisition system (B). Measurements were done through the opened window of the car with RGB, NIR, depth and thermal cameras as well as microphones. We recorded from the thermal camera at 50fps ((B), top center), the high framerate (HFR) camera with macro lens for face closeups at 120fps ((B), top right) and the high resolution camera in a stereo setup at 10fps ((B), top left). The stereo cameras covered the same field of view as the thermal camera. The camera setup was calibrated with a heated calibration board with circular pattern. Of the three possible lanes for the swab tests, the middle one was used during the time of our measurements.

**Fig. 2.**
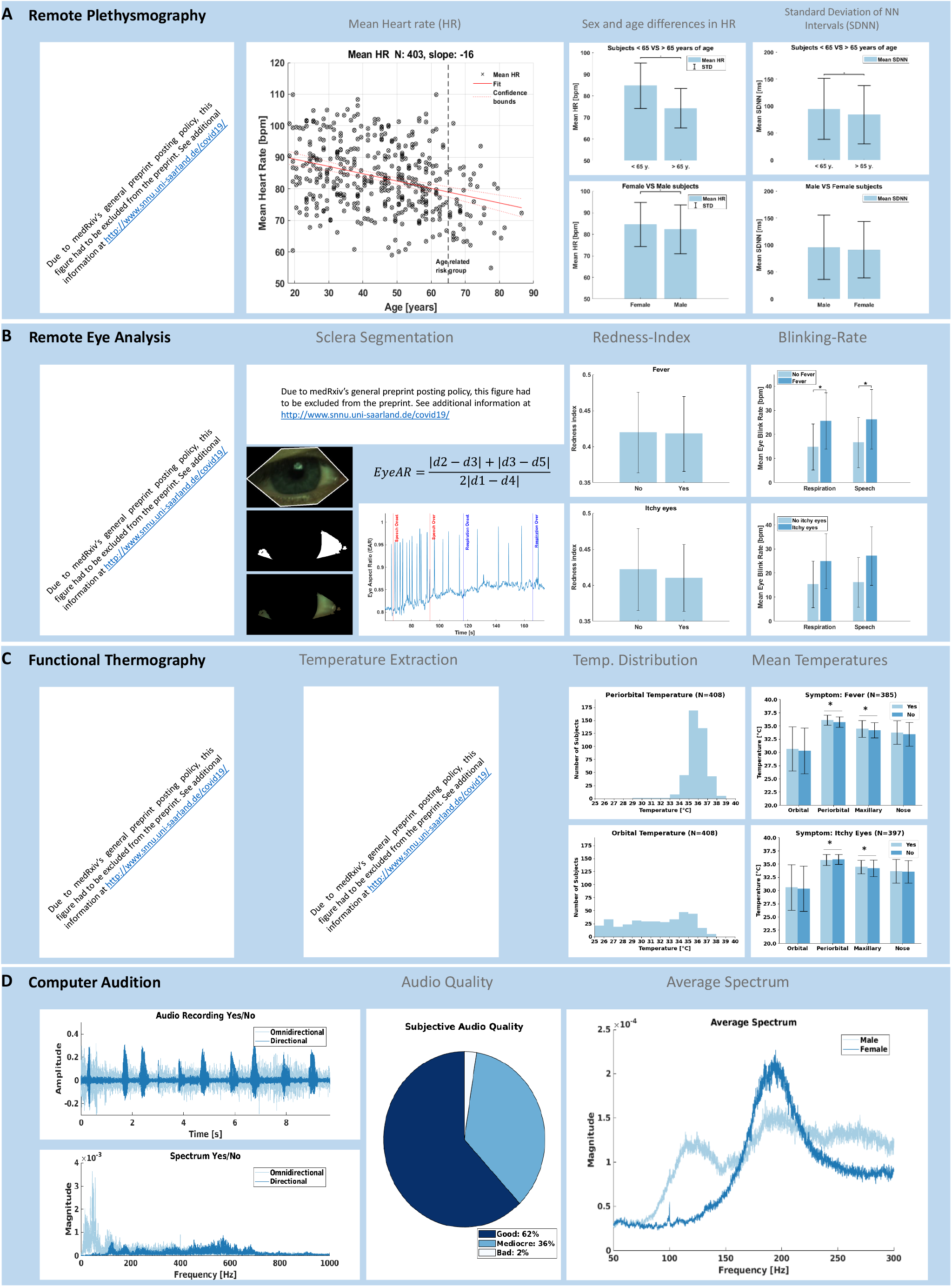
Results obtained in the reported evaluation period from the different modalities. rPPG (A) was used for the extraction of HR and HRV. For remote optometry (B) we analyzed EYE-AR and sclera redness. For static temperatures from the face (C), we looked at four points of interest. Manual assessment of our audio recordings (D) shows that 97.6% of our recordings had very good to acceptable sound quality, while for the rest the voice was rated unintelligible.

**Fig. 3.**
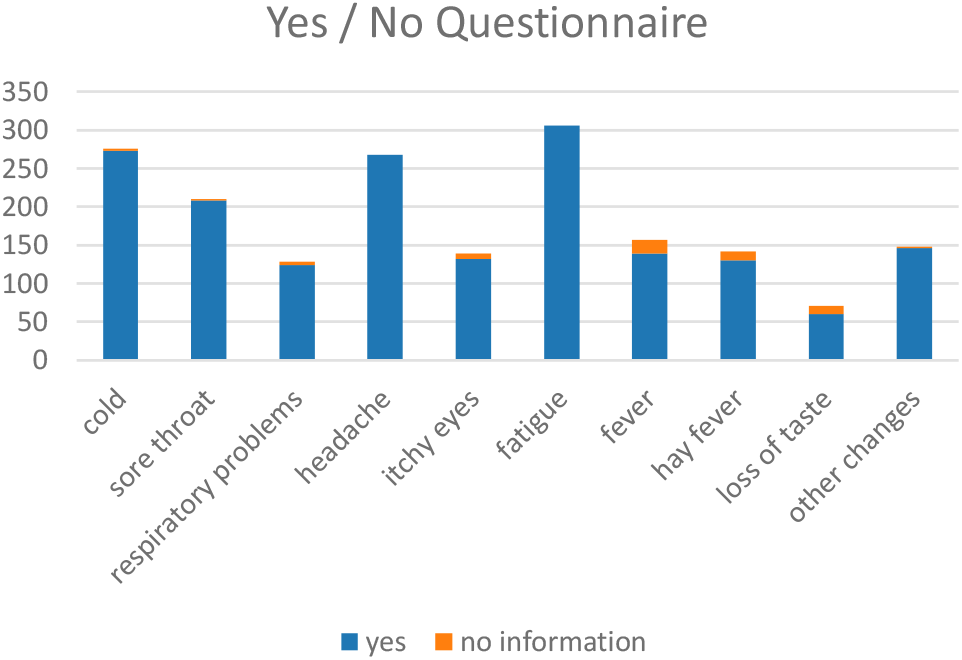
Counts of the symptoms reported by participants during the yes/no section of our experiment. Because every participant was already admitted to the DTSC based on the RKI recommendations regarding contact to infected persons and symptoms, most of our participants showed different flu-like symptoms or symptoms of a common cold. Most common were fatigue (306), followed by headache (268), cold (273), and sore throat (208).

## III. Results

We have recorded a dataset up to 6 minutes per subject with HFR or high resolution multimodal cameras and microphones. We applied already available procedures from computer vision and computer audition for assessment of the data quality. The evaluations and respective methodology used as proof of concept are described below. Fig. 2 summarizes the results for each of these modalities, in particular how the contactless physiological parameters compare to gender and age specific norm values and grouped them by a presence/absence of symptoms.

### A. Remote plethysmography

We applied the method of Wang et al. [10] for the extraction of remote plethysmography (rPPG) signals from skin segmented super-pixels of the HFR and 4k recordings and used custom scripts for peak detection and analysis. The analysis shows that the mean heart rate (MHR) decreases with age (see Fig. 2 (A)). Female participants show slightly higher values in MHR and lower values related to heart rate variability than male participants. This agrees with results from experiments with gold standard contact based sensors on large sample sizes [11], [12].

### B. Remote eye analysis

We precomputed landmarks for the HFR and 4k recordings using dlib [13]. On the eye, we analyzed the average blinking rate which can be a marker for drowsiness [14] and redness of the sclera as an indicator for follicular conjunctivitis [15] (see Fig. 2 (B)). We calculated the eye-aspect-ratio (Eye-AR) [16] from the pre-computed landmarks and apply custom algorithms to detect / count peaks for blinking detection. We found a significant difference in the blinking rate which was larger for participants reporting itchy eyes as compared to fever. We also extracted a region of interest around the eye and applied segmentation (gray scale based) to calculate the redness index (RI) from the sclera [15]. We found no significant difference of the RI between those with fever vs. itchy eyes.

### C. Functional Thermography

From the thermal recordings, we analyzed static temperatures from the orbital, periorbital, maxillary, and nose region (see Fig. 2 (C)). Vanilla landmark detectors did not perform consistently for all participants, so we applied a stack of preprocessing methods such as image inversion and unsharp masking to a set of 5 randomly sampled frames for each subject, applied a facial landmark detector [17] to each of the images with different pre-processing and averaged the results per frame. Failed detections were manually annotated. Our results showed a significant difference in temperature for subjects reporting fever vs. no fever in the maxillary, periorbital and nose region.

### D. Computer Audition

Schuller et al. argue how audio ‘in-the wild’ recordings under unconstrained conditions with various signal degradations can already have value for COVID19 computer audition [2]. With our platform, we get reproducible quality audio recordings from uniform hardware (see Fig 3 (D)). For instance, using established speech feature extraction schemes [18], the voice quality of our data is sufficient to solve a gender classification task with above 90% cross-validated accuracy of a support vector machine.

## IV. Discussion

A major limitation of our study is the marginal number of participants with positive PCR test result for SARS-CoV-2. The reason for this was the generally very low incidence rate in the study period in the region where the DTSC was located. In fact, the positive rate was below 0.4% at the DTSC Saarbrücken in the respective period. Thus, the specificity and sensitivity of the described approach with respect to SARS-CoV-2 infections cannot be assessed. However, our proof of concept shows that a remote data acquisition of SARS-CoV-2 infection related symptoms at DTSCs is possible. Our experiences and results enable the installation of similar approaches in regions that do massive DTSC testing.

Additionally, we have recorded an unprecedented, multi-modal dataset with a high number of subjects that can be used for the development and refinement of computer vision methods. Many state-of-the-art computer vision studies with isolated modalities have to resort to smaller, publicly available datasets: Considering the development of algorithms for the detection and classification of micro-expressions, Li et al. report between 80-210 subjects for common micro-expression datasets with evoked micro-expressions and they present a dataset with 20 subjects for spontaneous micro-expressions, recorded at 100fps (RGB) and 25fps (NIR) [19]. Davison et al. record 32 subjects at 200fps for spontaneous micro-expressions [20]. Our dataset contains recordings of 436 participants at 120fps (RGB), 50fps (thermal) and 30fps (NIR) over 6 minutes. While some subjects moved out of frame during various parts for the experiment for the HFR face close-up recordings, we expect a similar percentage of successful recordings that allow for micro-expression annotation as for our HR evaluations (see Fig. 2) and we can additionally report HFR thermal recordings with wider field of view.

For a facial landmarking task in the context of functional thermal imaging, datasets of around 2935 frames from 90 subjects with full manual annotation can be considered among the state-of-the-art [21]. Our 436 recordings at 50hz of up to 6 minutes potentially enable the generation of a dataset with 7-8 million frames which would be multiple orders of magnitudes larger. Our preliminary results suggest the possibility of partial, automatic annotation with an appropriate stack of pre-processing methods and together with tracking approaches could be used for a full annotation of the dataset in the future. On top of that, the setup potentially allows for multimodal mapping between NIR and thermal domain using the depth channel of the kinect and between RGB and thermal domain with the stereo setup (e.g. compare Palmero et al. [22]).

Computer vision algorithms require different conditions of environmental parameters. In the context of rPPG methods, Wang et al. require constant illumination to reconstruct PPG signals from videos in talking and static scenarios and various skin and then achieve high signal to noise ratio of the spectograms [10]. The employed studio illumination in our setup at the DTSC fulfills those requirements and allowed us to record high-quality data for rPPG measurements.

## V. Conclusion

We have proposed a setup to record multimodal data and have recorded a unique dataset of DTSC users across all age groups. To our knowledge, this is the first time that a multimodal video and audio dataset has been recorded at a SARS-CoV-2 DTSC. We have shown in our preliminary evaluations, that the data quality is sufficient for the application of already available procedures from computer vision and computer audition. This could allow for SARS-CoV-2 infection related symptoms assessment from such data in the future. While we captured only a marginal number of PCR positive subjects, our dataset can help with the development and refinement of computer vision algorithms beyond the COVID-19 pandemic: After full annotation for various computer vision tasks such as landmarking or micro-expression analysis, it has the potential to rank among the state-of-the-art in terms of the number of participants and age statistics. It has been recorded in an out-of-lab setting with realistic participant interaction, which for example could be encountered at the entrance of an employee parking area.

## Data Availability

See http://www.snnu.uni-saarland.de/covid19/ for information on data and source availability.

## Acknowledgments

We would like to thank the association of statutory health insurance physicians Saarland (KVSaar), in particular, Dr. Joachim Meiser and Michael Schneider for supporting the management with the local medical offices as well as for equipping us with PPE at the DTSC Saarbrücken. We thank the Federal Defense Forces of Germany, in particular Oberstleutnant Christoph Schacht, for their steady support during the study regarding the logistics and the installation of our data acquisition station at DTSC. We acknowledge the support of the ZF Friedrichshafen AG, in particular Florian Dauth, Volker Wagner, and Dr. Peter Reitz for supporting us with the research vehicle used for the data acquisition. A special note of thanks goes to the responsible authorities at the Regionalverband (regional association of towns) Saarbrücken, in particular Peter Gillo and Alexander Birk and the responsible ethics commission at the medical council Saarland for helping us to resolve all the data security and ethical issues in an incredible short amount of time. Furthermore, the authors would like to thank Benedikt Buchheit, Maximilian Becker, Dr. Farah I. Corona-Strauss, Adrian Mai, Richard Morsch, Patrick Schäfer, and Elena Schneider from our research unit for supporting the administration as well as the data processing. Finally, we would like to thank Professor Alexander L. Francis from Purdue University for proving valuable feedback on the first version of this manuscript.

## References

[1] A. S. Manolis, A. A. Manolis, T. A. Manolis, E. J. Apostolopoulos, D. Papatheou, and H. Melita, “COVID-19 infection and cardiac arrhythmias,” Trends in Cardiovascular Medicine, aug 2020.

[2] B. W. Schuller, D. M. Schuller, K. Qian, J. Liu, H. Zheng, and X. Li, “COVID-19 and Computer Audition: An Overview on What Speech & Sound Analysis Could Contribute in the SARS-CoV-2 Corona Crisis,” mar 2020.

[3] B. J. Quilty, S. Clifford, S. Flasche, R. M. Eggo, and Others, “Effectiveness of airport screening at detecting travellers infected with novel coronavirus (2019-nCoV),” Eurosurveillance, vol. 25, no. 5, pp. 2000080, 2020.

[4] Y.-F. Tu, C.-S. Chien, A. A. Yarmishyn, Y.-Y. Lin, Y.-H. Luo, Y.-T. Lin, W.-Y. Lai, D.-M. Yang, S.-J. Chou, Y.-P. Yang, and Others, “A review of SARS-CoV-2 and the ongoing clinical trials,” International journal of molecular sciences, vol. 21, no. 7, pp. 2657, 2020.

[5] R. Wölfel, V. M. Corman, W. Guggemos, M. Seilmaier, S. Zange, M. A. Müller, D. Niemeyer, T. C. Jones, P. Vollmar, C. Rothe, and Others, “Virological assessment of hospitalized patients with COVID-2019,” Nature, vol. 581, no. 7809, pp. 465–469, 2020.

[6] W.-j. Guan, Z.-y. Ni, Y Hu, W.-h. Liang, C.-q. Ou, J.-x. He, L. Liu, H Shan, C.-l. Lei, D. S. C. Hui, and Others, “Clinical characteristics of coronavirus disease 2019 in China,” New England journal of medicine, vol. 382, no. 18, pp. 1708–1720, 2020.

[7] J.-P. O. Li, D. S. C. Lam, Y. Chen, and D. S. W. Ting, “Novel Coronavirus disease 2019 (COVID-19): The importance of recognising possible early ocular manifestation and using protective eyewear,” 2020.

[8] K. T. Kwon, J.-H. Ko, H. Shin, M. Sung, and J. Y. Kim, “Drive-through screening center for covid-19: A safe and efficient screening system against massive community outbreak,” Journal of Korean Medical Science, vol. 35, no. 11, 2020.

[9] Robert Koch-Institut, “COVID-19-Verdacht: Maßnahmen und Testkriterien - Orientierungshilfe für Ä rzte,” 2020.

[10] W. Wang, A. C. Den Brinker, S. Stuijk, and G. De Haan, “Algorithmic Principles of Remote PPG,” IEEE Transactions on Biomedical Engineering, vol. 64, no. 7, pp. 1479–1491, 2017.

[11] R. Zhao, D. Li, P. Zuo, R. Bai, Q. Zhou, J. Fan, C. Li, L. Wang, and X. Yang, “Influences of Age, Gender, and Circadian Rhythm on Deceleration Capacity in Subjects without Evident Heart Diseases,” Annals of Noninvasive Electrocardiology, vol. 20, no. 2, pp. 158–166, mar 2015.

[12] J. Koenig and J. F. Thayer, “Sex differences in healthy human heart rate variability: A meta-analysis,” Neuroscience & Biobehavioral Reviews, vol. 64, may 2016.

[13] D. E. King, “Dlib-ml: A machine learning toolkit,” Journal of Machine Learning Research, vol. 10, 2009.

[14] R. Martins and J. M. Carvalho, “Eye blinking as an indicator of fatigue and mental load—a systematic review,” in Occupational Safety and Hygiene III. 2015.

[15] I. Sárándi, D. P. Claßen, A. Astvatsatourov, O. Pfaar, L. Klimek, R. Mösges, and T. M. Deserno, “Quantitative conjunctival provocation test for controlled clinical trials,” Methods of Information in Medicine, vol. 53, no. 4, pp. 238–244, 2014.

[16] J. Cech and T. Soukupova, “Real-Time Eye Blink Detection using Facial Landmarks,” Center for Machine Perception, Department of Cybernetics Faculty of Electrical Engineering, Czech Technical University in Prague, 2016.

[17] A. Bulat and G. Tzimiropoulos, “How far are we from solving the 2d & 3d face alignment problem?(and a dataset of 230,000 3d facial landmarks),” in Proceedings of the IEEE International Conference on Computer Vision, 2017, pp. 1021–1030.

[18] F. Eyben, M. Wöllmer, and B. Schuller, “OpenSMILE - The Munich versatile and fast open-source audio feature extractor,” MM’10 - Proceedings of the ACM Multimedia 2010 International Conference, pp. 1459–1462, 2010.

[19] X. Li, T. Pfister, X. Huang, G. Zhao, and M. Pietikäinen, “A spontaneous micro-expression database: Inducement, collection and baseline,” in 2013 10th IEEE International Conference and Workshops on Automatic face and gesture recognition (fg). IEEE, 2013, pp. 1–6.

[20] A. K. Davison, C. Lansley, N. Costen, K. Tan, and M. H. Yap, “Samm: A spontaneous micro-facial movement dataset,” IEEE Transactions on Affective Computing, vol. 9, no. 1, pp. 116–129, 2016.

[21] M. Kopaczka, R. Kolk, J. Schock, F. Burkhard, and D. Merhof, “A thermal infrared face database with facial landmarks and emotion labels,” IEEE Transactions on Instrumentation and Measurement, vol. 68, no. 5, pp. 1389–1401, 2018.

[22] C. Palmero, A. Clapés, C. Bahnsen, A. Møgelmose, T. B. Moeslund, and S. Escalera, “Multi-modal rgb–depth–thermal human body segmentation,” International Journal of Computer Vision, vol. 118, no. 2, pp. 217–239, 2016.

